# Left Atrial Appendage Closure, Direct Oral Anticoagulants or Warfarin in Atrial Fibrillation: A Systematic Review and Network Meta-analysis of Randomized Clinical Trials

**DOI:** 10.64898/2026.05.07.26352700

**Authors:** Samir B. Pancholy, M. Haisum Maqsood, Muhammad Sabih Saleem, Dipen Zalavadia, Khyati Khattar, Tejas M. Patel, Sripal Bangalore

## Abstract

**Background:** Left atrial appendage closure (LAAC) and direct oral anticoagulants (DOACs) have emerged as alternatives to warfarin for stroke prevention in atrial fibrillation (AF). However, recent trials have shown variable results igniting the debate on this topic.

**Methods:** We performed a systematic review and network meta-analysis (NMA) of RCTs comparing LAAC, DOACs, and warfarin in patients with AF. The primary efficacy outcome was ischemic stroke or systemic embolism (IS/SE) and the primary bleeding outcome was hemorrhagic stroke (HS). Secondary outcomes included net adverse clinical events (NACE) and major or clinically relevant bleeding (MCRB). Pooled odds ratios (ORs) with 95% confidence intervals (CIs) were estimated using a random-effects model.

**Results:** Ten RCTs (LAAC: 6 trials; DOAC: 8 trials; warfarin: 6 trials) enrolling 78,594 patients fulfilled the inclusion criteria. There were no significant differences for the primary efficacy outcome of IS/SE among the 3 strategies. However, when compared with warfarin, both DOACs (OR 0·43, 95% CI 0·34–0·54) and LAAC (OR 0·34, 95% CI 0·18–0·63) reduced the primary safety outcome of HS, with no significant difference between them (OR 0·79, 95% CI 0·44–1·3). For NACE, both DOACs (OR 0·87, 95% CI 0·83–0·91) and LAAC (OR 0·85, 95% CI 0·73–0·99) were superior to warfarin, with similar performance between them (OR 0·98, 95% CI 0·84–1·13). For MCRB, DOACs were superior to warfarin (OR 0·79, 95% CI 0·63–0·99), while LAAC showed a non-significant trend towards benefit.

**Conclusion:** In this meta-analysis of RCTs with data from over 78,000 patients, LAAC and DOACs significantly reduced NACE driven by lower hemorrhagic stroke and provided equivalent IS/SE protection compared with warfarin, making LAAC a potential viable alternative to oral anticoagulation in appropriately selected AF patients.

**Funding:** None.

## INTRODUCTION

Atrial fibrillation (AF) affects more than 37 million individuals worldwide and represents the most prevalent sustained cardiac arrhythmia encountered in clinical practice (1). Cardioembolic stroke predominantly arising from thrombus formation within the left atrial appendage (LAA) is the most feared complication of AF and is associated with substantial morbidity and mortality (2). Vitamin K antagonists (VKAs), exemplified by warfarin, served as the cornerstone of stroke prophylaxis in AF for decades. However, the narrow therapeutic index of warfarin, obligatory international normalized ratio monitoring, dietary interactions, and its associated hemorrhagic burden, particularly intracranial bleeding prompted the search for safer, more convenient alternatives.

The advent of direct oral anticoagulants (DOACs) have transformed AF management. Landmark phase III randomized clinical trials (RCTs) demonstrated non-inferiority or superiority of DOACs versus warfarin for stroke prevention, with consistently lower rates of intracranial hemorrhage and improved pharmacokinetic predictability (5–8). Despite these advantages, DOACs require sustained adherence and remain hazardous in patients with severe renal impairment, prior major bleeding, or complex polypharmacy.

Left atrial appendage closure (LAAC), offers a non-pharmacological, catheter-based approach to stroke prevention by mechanically excluding the LAA, the anatomic source of >90% of AF-related thrombi (10). Early trials established non-inferiority of LAAC versus warfarin for the composite of stroke, systemic embolism, and cardiovascular death, with a progressive hemorrhagic benefit over long-term follow-up (11,12). More recently, the Closure-AF and CHAMPION-AF trials comparing LAAC with DOACs across diverse patient populations reached opposite conclusions, igniting a debate on the efficacy and safety of these devices (15–17).

Accordingly, we performed a systematic review and meta-analysis of all available RCTs comparing LAAC, DOACs, and warfarin to assess the relative efficacy and safety in patients with AF.

## METHODS

### Study Design and Registration

This systematic review and NMA were conducted in accordance with the Preferred Reporting Items for Systematic Reviews and Meta-Analyses extension for Network Meta-Analyses (PRISMA-NMA) guidelines (18). The protocol was prospectively registered with PROSPERO registry (ID: CRD420261362040). The data supporting the results of this study will be made available from the corresponding author upon reasonable request. As this was a trial level meta-analysis of published clinical data, institutional review board evaluation was not deemed necessary.

### Search Strategy and Eligibility Criteria

A systematic search of PubMed/MEDLINE, EMBASE, and the Cochrane Central Register of Controlled Trials (CENTRAL) was conducted from inception through 04/08/2026. Medical Education Subject Headings (MeSH) included combinations of ’atrial fibrillation’, ’left atrial appendage closure’, ’Watchman’, ’direct oral anticoagulant’, ’DOAC’, ’NOAC’, ’warfarin’, ’randomized controlled trial’, and related MeSH terms. Conference proceedings from the American College of Cardiology (ACC), European Society of Cardiology (ESC), American Heart Association (AHA), and Trans catheter Cardiovascular Therapeutics (TCT) annual meetings, as well as ClinicalTrials.gov and FDA.gov were additionally searched to minimize publication bias. There was no language restriction for the search.

Eligible studies were RCTs that (1) enrolled adults with AF without mitral stenosis or mechanical prosthetic heart valves; (2) directly compared at least two of the three interventions (LAAC, DOAC, and warfarin); and (3) reported at least one pre-specified outcome. Studies enrolling only non-AF populations, observational designs, or evaluating non-regulatory-approved device platforms were excluded.

### Data Extraction and Outcomes

Two reviewers (SBP, MHM) independently extracted data using a standardized template. Primary efficacy outcome was IS/SE while primary safety outcome was hemorrhagic stroke. Secondary outcomes included Net Adverse Clinical Events (NACE), defined as a composite of ischemic stroke, systemic embolism, cardiovascular death, major bleeding, and major or clinically relevant non-major bleeding. Outcome definitions were harmonized across trials using published adjudication criteria.

The treatment interventions were categorized into 3 groups: 1) LAAC; 2) DOAC; and 3) warfarin (reference treatment arm).

### Risk of Bias and Certainty of Evidence

Risk of bias was assessed using the Cochrane Risk of Bias 2.0 (RoB2) tool across five domains: randomization process, deviations from intended interventions, missing outcome data, outcome measurement, and selection of reported results. The GRADE framework was applied to rate the overall certainty of evidence for each network estimate.

### Statistical Analysis

We reported continuous variables as means with standard deviations and categorical variables as frequencies with percentages. For efficacy and safety outcomes, adverse events using odds ratio (OR) with 95% confidence intervals (CI) were calculated. Frequentist estimation of network meta-analysis model was used for LAAC, DOAC, and warfarin (reference treatment arm). Effect size of each study and treatment were evaluated using interval plot with warfarin as the reference treatment arm (19–22). We also performed other pairs of comparisons. We ranked ordered the treatment strategies using surface under the cumulative ranking (SUCRA) visualized using the rankograms (23). We evaluated publication bias using network funnel plot (24,25). We evaluated inconsistency between the trials at network level (24). A two-sided p-values <0.05 was considered for statistical significance. All analyses were performed in the intention-to-treat population. Stata version 19.5 software (Stata Corporation, College Station, TX, USA) with mvmeta package was used for the analyses.

## RESULTS

### Study Selection and Characteristics

The systematic search identified 2202 records; after removing duplicates and screening the titles, 10 RCTs met eligibility criteria. Trial characteristics are summarized in **Table 1**. The network included three treatment nodes: LAAC (6 trials), DOAC (8 trials), and warfarin (6 trials) (5–8, 11–13, 15–17). Enrolled patients had a mean weighted age of 71.7 years and a mean weighted CHA₂DS₂-VASc score of 2.7 reflecting a high-risk AF population. Mean follow-up ranged from 1.0 to 3.5 years. Device-based trials were open-label by necessity; pharmacological trials employed double-blind designs where feasible. The risk of bias was low to moderate across the included trials.

**Table 1.** Characteristics of included randomized clinical trials.

| **Trial** | **Year** | **Comparators** | **N** | **Age**<br>**(Years)** | **CHA<sub>2</sub>DS<sub>2</sub>-VASc**<br>**Score** | **Follow-up** | **Outcomes** |
| --- | --- | --- | --- | --- | --- | --- | --- |
| PROTECT AF | 2009 | LAAC vs warfarin | 707 | 71.7 | 3.4 | 2.3 years | IS/SE, HS, NACE,<br>MCRB |
| PREVAIL | 2014 | LAAC vs warfarin | 407 | 74.0 | 3.8 | 1year | IS/SE, HS, NACE,<br>MCRB |
| Prague-17 | 2020 | LAAC vs DOAC | 402 | 73.3 | 4.7 | 3.5 years | IS/SE, HS, NACE,<br>MCRB |
| Closure-AF | 2024 | LAAC vs DOAC | 750 | 75.0 | 4.5 | 2 years | IS/SE, HS, NACE,<br>MCRB |
| OPTION | 2023 | LAAC vs DOAC | 180 | 73.8 | 4.2 | 1 year | IS/SE, HS |
| CHAMPION-AF | 2024 | LAAC vs DOAC | 3,000* | 74.5 | 4.6 | 2.5 years | IS/SE, HS, NACE,<br>MCRB |
| RE-LY | 2009 | DOAC vs warfarin | 18,113 | 71.5 | 3.5 | 2 years | IS/SE, HS, MCRB |
| ROCKET-AF | 2011 | DOAC vs warfarin | 14,264 | 73.0 | 3.5 | 1.9 years | IS/SE, HS, MCRB |
| ARISTOTLE | 2011 | DOAC vs warfarin | 18,201 | 70.0 | 3.5 | 1.8 years | IS/SE, HS, MCRB |
| ENGAGE AF | 2013 | DOAC vs warfarin | 21,105 | 72.0 | 4.2 | 2.8 years | IS/SE, HS, MCRB |

IS/SE indicates ischemic stroke or systemic embolism; HS, hemorrhagic stroke; NACE, net adverse clinical events; MCRB, major bleeding or clinically relevant; DOAC, direct oral anticoagulant; LAAC, left atrial appendage closure. \*Estimated enrollment.

**Table 2.** Network meta-analysis results summary.OR indicates odds ratio; CI, confidence interval; IS/SE, ischemic stroke or systemic embolism; NACE, net adverse clinical events; NS, not significant. Bold values indicate statistical significance (95% CI excludes unity).

| Outcome | Comparison | OR (95% CI) | Interpretation |
| --- | --- | --- | --- |
| Ischemic stroke/SE | DOAC vs warfarin | 0.95 (0.79–1.15) | No significant difference |
|  | LAAC vs warfarin | 1.02 (0.72–1.45) | No significant difference |
|  | LAAC vs DOAC | 1.07 (0.78–1.47) | No significant difference |
| NACE | DOAC vs warfarin | 0.87 (0.83–0.91) | DOAC superior to warfarin |
|  | LAAC vs warfarin | 0.85 (0.73–0.99) | LAAC superior to warfarin |
|  | LAAC vs DOAC | 0.98 (0.84–1.13) | No significant difference |
| Hemorrhagic stroke | DOAC vs warfarin | 0.43 (0.34–0.54) | DOAC superior to warfarin |
|  | LAAC vs warfarin | 0.34 (0.18–0.63) | LAAC superior to warfarin |
|  | LAAC vs DOAC | 0.79 (0.44–1.43) | No significant difference |
| Major/clinically relevant bleeding | DOAC vs warfarin | 0.79 (0.63–0.99) | DOAC superior to warfarin |
|  | LAAC vs warfarin | 0.72 (0.50–1.03) | Trend favoring LAAC (NS) |
|  | LAAC vs DOAC | 0.91 (0.68–1.20) | No significant difference |

### Ischemic stroke or systemic embolism

Ten RCTs enrolling 78,594 patients provided data for this outcome (LAAC = 6 trials with 3681 patients, DOAC = 8 trials with 45302 patients, and Warfarin = 6 trials with 29611 patients) (**Figure 1 panel A, Supplement Figure S2 panel A**). There was no significant difference in ischemic stroke or systemic emboli with DOAC or LAAC compared with warfarin **(Figure 1 panel B)**. The outcome between DOAC and LAAC was similar **(Figure 1 panel B)**. There was no evidence of inconsistency in the network meta-analysis (p=0.38) or presence of publication bias **(Supplement Figure S2 panel C)**.

**Figure 1.**
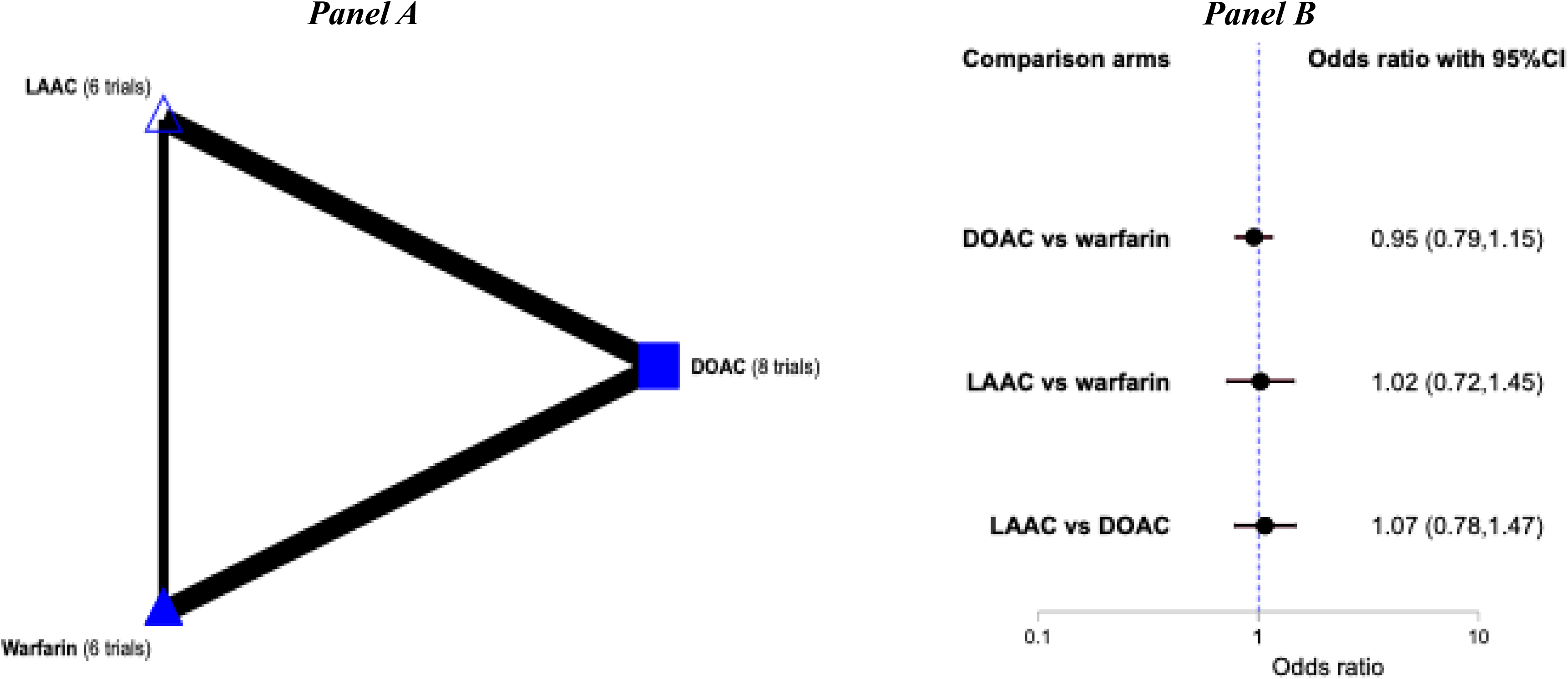
Primary Efficacy Outcome of Ischemic Stroke or Systemic Emboli. Panel A: Network map. Nodes represents comparison arms. Thickness of lines represents number of trials for the comparison. **Panel B: Interval plot comparing the three treatment strategies.** Primary analysis compares each treatment arm with warfarin as reference. *Abbreviations: LAAC – left atrial appendage closure; DOAC – direct oral anti-coagulants.

### Net adverse clinical events (NACE)

Seven RCTs enrolling 63,169 patients provided data for this outcome (LAAC = 4 trials with 2949 patients, DOAC = 7 trials with 38,134 patients, and Warfarin = 3 trials with 22,086 patients) (**Figure 2 panel A, Supplement Figure S4 panel A**). There was a lower risk of NACE with DOAC (OR=0·87; 95% CI: 0·83–0·91) and LAAC (OR=0·85; 95% CI: 0·73–0·99) when compared with warfarin **(Figure 2 panel B)**. There were no significant differences in NACE between LAAC and DOAC **(Figure 2 panel B)**. LAAC ranked #1, followed by DOAC (#2), and warfarin (#3) **(Supplement Figure S4 panel B)**. There was no evidence of publication bias **(Supplement Figure S4 panel C)**, but there was evidence for inconsistency (p<0.001) in the overall model. Further evaluation did not show inconsistency at all three comparison arms (LAAC vs DOAC and DOAC vs Warfarin) (**Supplement Table S3**). There was no loop in this network so the cause of inconsistency was not further evaluated.

**Figure 2.**
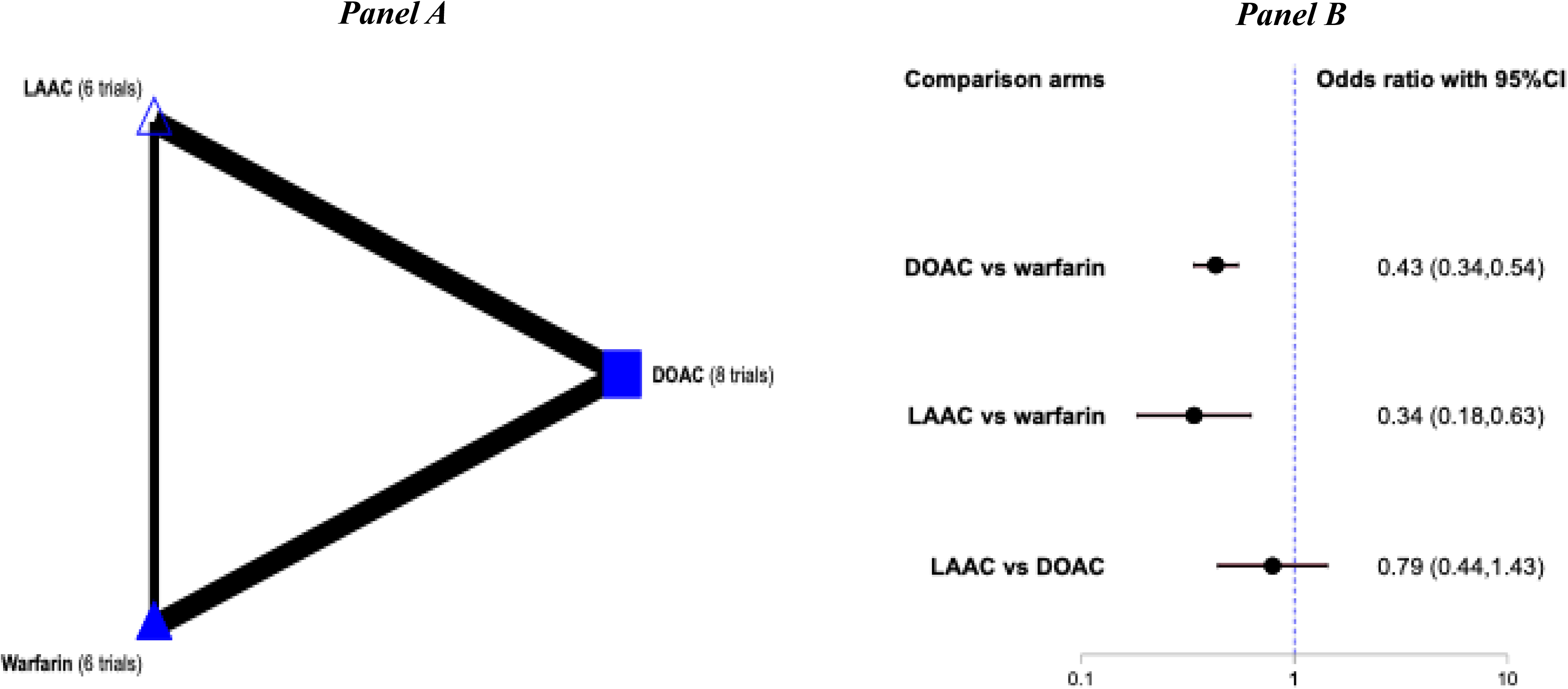
Secondary Outcome of Net Adverse Clinical Events. Panel A: Network map. Nodes represents comparison arms. Thickness of lines represents number of trials for the comparison. **Panel B: Interval plot comparing the three treatment strategies.** Primary analysis compares each treatment arm with warfarin as reference. *Abbreviations: LAAC – left atrial appendage closure; DOAC – direct oral anti-coagulants.

**Figure 3.**
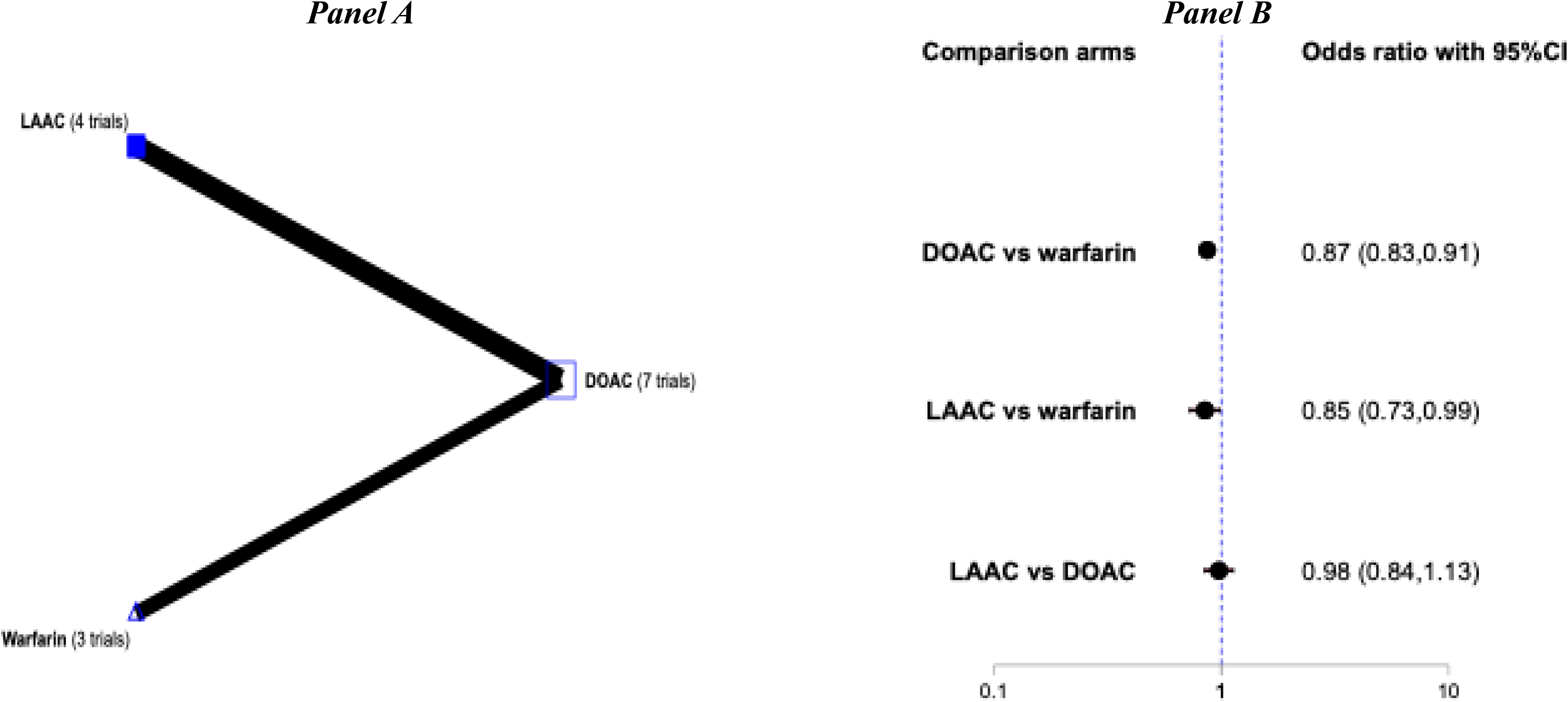
Primary safety Outcome of Hemorrhagic stroke. Panel A: Network map. Nodes represents comparison arms. Thickness of lines represents number of trials for the comparison. **Panel B: Interval plot comparing the three treatment strategies.** Primary analysis compares each treatment arm with warfarin as reference. *Abbreviations: LAAC – left atrial appendage closure; DOAC – direct oral anti-coagulants.

### Major or clinically relevant bleeding

Seven RCTs enrolling 59,292 patients provided data for this outcome (LAAC = 4 trials with 2949 patients, DOAC = 7 trials with 33,154 patients, and Warfarin = 3 trials with 23,189 patients) (**Figure 4 panel A, Supplement Figure S5 panel A**). There was a lower risk of major or clinically relevant bleeding with DOAC (OR=0·79; 95% CI: 0·63–0·99) compared with warfarin **(Figure 4 panel B)**. There were no significant differences in other comparison arms **(Figure 4 panel B)**. DOAC ranked #1, followed by LAAC (#2), and warfarin (#3) **(Figure 2 panel C, Supplement Figure S5 panel B)**. There was no evidence of publication bias **(Supplement Figure S5 panel C)**, but there was evidence for inconsistency (p=0·042) in the overall model. Further evaluation did not show inconsistency at all three comparison arms (LAAC vs DOAC and DOAC vs Warfarin) (**Supplement Table S4**). There was no loop in this network so the cause of inconsistency was not further evaluated.

**Figure 4.**
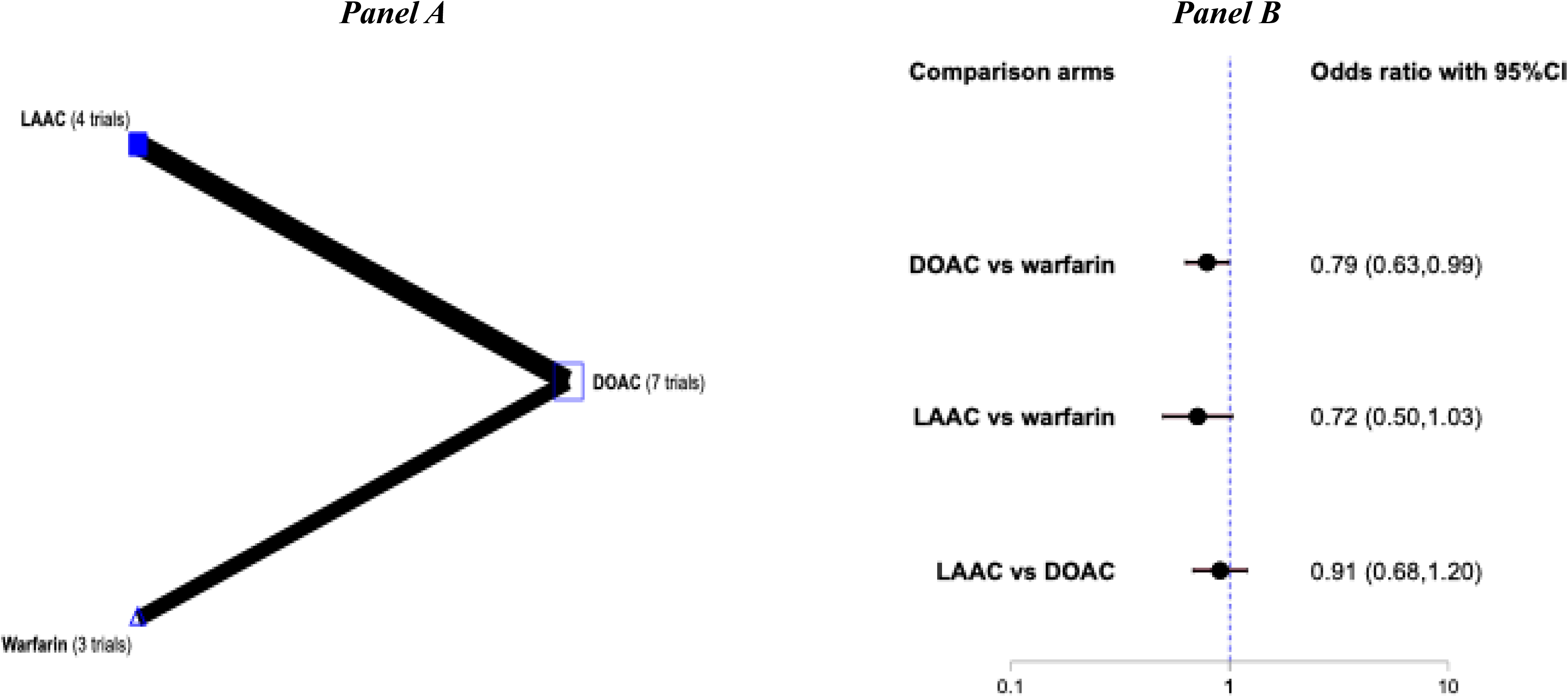
Outcomes of Major or Clinically Relevant Bleeding. Panel A: Network map. Nodes represents comparison arms. Thickness of lines represents number of trials for the comparison. **Panel B: Interval plot comparing the three treatment strategies.** Primary analysis compares each treatment arm with warfarin as reference. *Abbreviations: LAAC – left atrial appendage closure; DOAC – direct oral anti-coagulants.

## DISCUSSION

In this NMA of 10 RCTs with over 78,000 patients, DOACs and LAAC reduced NACE, driven by lower hemorrhagic stroke and lower major or clinically relevant bleeding with similar protection against ischemic stroke or systemic emboli when compared with warfarin.

The results of this meta-analysis showing similar outcomes of LAAC, DOACs, and warfarin for IS/SE is a clinically critical observation. It directly addresses the longstanding concern that LAAC, by eliminating systemic anticoagulation rather than supplementing it, might incompletely protect against all cardioembolic mechanisms. The current NMA, leveraging the combined precision of 10 RCTs, provides the strongest available evidence that LAAC achieves similar ischemic stroke and embolism protection relative to both warfarin and contemporary DOACs, a conclusion consistent with the pre-specified non-inferiority findings of PROTECT AF, PREVAIL, and Prague-17, but now formally established through a simultaneous three-node network.

The hemorrhagic stroke findings deserve particular emphasis. Warfarin’s intracranial hemorrhage burden represents its most clinically devastating limitation, with fatality rates exceeding 50% among affected patients (25). Our analysis confirms that this risk is substantially mitigated by either DOACs or LAAC. The confidence intervals for LAAC versus DOACs for HS cross unity (OR 0·79, 0·44–1·43), precluding definitive superiority claims; however, the directionality and upper CI bound of 1·43 are reassuring regarding LAAC’s hemorrhagic safety profile. These findings align with the biological rationale of LAAC: once the device endothelializes and potent antiplatelet therapy is discontinued, the patient is no longer exposed to anticoagulant mediated bleeding risk, which accumulates progressively over years of pharmacotherapy, a benefit documented in long-term follow-up of PROTECT AF.

Notably, for NACE and major bleeding outcomes, there was no direct comparison of LAAC and warfarin, and the estimates rely on indirect evidence. This methodological feature, reflected in 50% indirect network weighting, appropriately tempers the certainty of evidence for LAAC-warfarin NACE and bleeding comparisons (GRADE: moderate). Nonetheless, the LAAC vs DOAC NACE estimate (OR 0·98, 0·84–1·13) is noteworthy for its near-unity point estimate and narrow CI, providing strong evidence of clinical equivalence between these two stroke prevention strategies for composite net outcomes, a direct replication of Prague-17’s central conclusion in a larger pooled framework.

The rankograms provide additional insight into the probabilistic treatment hierarchy. For HS, LAAC had the highest probability of ranking best, while for IS/SE all three treatments were comparably ranked. For major bleeding, LAAC ranked highest, followed by DOACs, with warfarin consistently last. These findings are consistent with contemporary data from the PINNACLE FLX registry, which demonstrates the safety profile of the next-generation Watchman FLX device (26). Collectively, the ranking data support a narrative in which LAAC may offer the most favorable overall bleeding profile among available strategies, albeit with the tradeoff of a procedural risk (∼2–4% for major device-related adverse events in contemporary series) that must be individualized based on patient preference, bleeding history, and anticoagulant adherence.

Several limitations merit consideration. First, this is a trial level meta-analysis with limited granularity and hence temporal associations, and additional analyses could not be performed. Second, the included DOAC trials span nearly two decades, with evolving background therapies, patient selection criteria, and event adjudication methodologies. While the design-by-treatment interaction tests support global network consistency, heterogeneity in warfarin-arm event rates across LAAC and DOAC trials remains a potential source of between-study variance. Third, LAAC trials enrolled populations with higher HAS-BLED scores and greater prevalence of anticoagulation contraindications, creating potential effect modification, that aggregate NMA estimates cannot fully decompose without individual participant data. Fourth, follow-up varied substantially (1·0–3·5 years); the cumulative hemorrhagic benefit of LAAC, which accrues progressively after antiplatelet discontinuation, may be underestimated in shorter-duration RCTs. Fifth, device generation (Watchman FLX vs. original Watchman vs. Amulet) was heterogeneous across trials, and platform-specific subgroup analyses were unavailable within the aggregate framework. Sixth, the NACE and major bleeding networks lacked a direct LAAC vs. warfarin comparison, limiting inference for those comparisons to indirect estimates.

In conclusion, this NMA of 10 RCTs demonstrates that LAAC and DOACs are both safer for NACE and hemorrhagic stroke/major bleeding compared to warfarin in patients with AF. The outcomes with LAAC and DOACs were largely similar to each other and at a minimum similar to that of warfarin for ischemic stroke prevention in AF. These findings support positioning LAAC as an evidence-based alternative to indefinite oral anticoagulation in appropriately selected patients with non-valvular AF, particularly those with a history of, or elevated risk for anticoagulant-related bleeding, demonstrated poor adherence, or patient preference for a non-pharmacological approach. Future RCTs with longer follow-up, standardized NACE definitions, and stratification by device generation will further refine these estimates.

## Disclosures

Dr. Bangalore: Advisory board/consultant for Abbott Vascular, Boston Scientific, Shockwave, Stryker, AngioDynamics, Imperative Care and Jupiter.

The rest of the authors have no conflicts of interest to declare.

## Funding

None.

## Data Availability

Given the fact that this is a meta-analysis of trial level data, the data we have will be available upon request.Given the fact that this is a meta-analysis of trial level data, the data we have will be available upon request.

## Notes

### Competing Interest Statement

The authors have declared no competing interest.

### Clinical Trial

PROSPERO registry (ID: CRD420261362040)

### Funding Statement

No extramural funding was available for this project.

### Author Declarations

This was a trial level, meta-analysis, and so no institutional review. Board approval was deemed necessary

